# Adolescent vaccination with BNT162b2 (Comirnaty, Pfizer-BioNTech) vaccine and effectiveness against COVID-19: national test-negative case-control study, England

**DOI:** 10.1101/2021.12.10.21267408

**Authors:** Annabel A Powell, Freja Kirsebom, Julia Stowe, Kelsey McOwat, Vanessa Saliba, Mary E Ramsay, Jamie Lopez-Bernal, Nick Andrews, Shamez N Ladhani

## Abstract

Adolescents in the UK were recommended to have their first dose of mRNA vaccine during a period of high community transmission due to the highly transmissible Delta variant, followed by a second dose at an extended interval of 8-12 weeks. We used national SARS-CoV-2 testing, vaccination and hospitalisation data to estimate vaccine effectiveness (VE) using a test-negative case-control design, against PCR-confirmed symptomatic COVID-19 in England. BNT162b2 vaccination in 12-15-year-olds and 16-17-year-olds was associated with lower VE against symptomatic COVID-19 caused by Omicron compared to Delta. Data shows a rapid increase in VE against symptomatic COVID-19 after the second dose for both Delta and Omicron, although this declines to 23% against Omicron after 70+ days. Very high protection was achieved for Delta against hospitalisation after one dose. Our data highlight the importance of the second vaccine dose for protection against symptomatic COVID-19 and raise important questions about the objectives of an adolescent immunisation programme. If prevention of infection is the primary aim, then regular COVID-19 vaccine boosters will be required.

## Background

Children and adolescents have a low risk of severe COVID-19.^1^ In the UK, COVID-19 vaccination for adults began in December 2020.^2^ Because of concerns about rare but potentially severe myocarditis after mRNA vaccination, mainly after the second dose in young adult males, the UK Joint Committee on Vaccination and Immunisation (JCVI) initially recommended one dose for 16-17-year-olds from 4 August 2021,^3^ and recommended against vaccinating 12-15-year-olds because of marginal risk-benefits,^4^ although UK ministers subsequently recommended vaccinating this group with BNT162b2 (Comirnaty, Pfizer-BioNTech) or mRNA-1273 (Spikevax, Moderna) from 13 September 2021 to prevent education disruption.^5^ Contrary to the authorised 3-week interval, the UK recommends 8-12 weeks between doses, because of the high protection the first dose provides and higher antibody responses after a later second dose.^6^ The UK strategy provided a unique opportunity to assess single-dose mRNA vaccine effectiveness (VE) in adolescents during a period of high community infection with the highly-transmissible Delta variant and, subsequently with the more-transmissible and now dominant Omicron variant^7^.

## Methods

We used a test-negative case-control design to estimate VE after one BNT162b2 dose against PCR-confirmed symptomatic COVID-19 with the Delta and Omicron variants in England, as described previously.^8,9^ Vaccination status in symptomatic 12-15-year-olds and 16-17-year-olds with PCR-confirmed SARS-COV-2 infection was compared with vaccination status in symptomatic adolescents in the same age-groups who had a negative SARS-COV-2 PCR test. (full details in **Supplement 1)**.

## Results

From week 37 2021 onwards, there were 617,259 eligible tests for 12-15-year-olds and 225,670 for 16-17-year-olds, with a test date within 10 days of symptom onset date, and which could be linked to the National Immunisation Management system (match rate: 92.5%) (**Supplement 3&4**). Vaccine uptake and confirmed infections by age-group and over time are summarised in **Supplement 5**.

After one vaccine dose in 12-15-year-olds VE against symptomatic disease with Delta after dose one peaked at days 14-20 after vaccination (74.5%; 95%CI 73.2-75.6) and then declined gradually, reaching 45.9% (95%CI 41.2-50.1) at days 70-83 post-vaccination **(Figure 1, Supplement 6)**. For Omicron, VE was significantly lower at these timepoints, peaking at 49.6% (95CI% 43.9-54.8) and dropping to 16.1% (95CI% 12.1-20.0), respectively. After two doses, VE increased and peaked 7-13 days later at 93.2% (95%CI 81.5-97.5) for Delta and at 83.1% (95%CI 78.2-86.9) for Omicron. (**Supplement 6**) For Delta, VE against hospitalisation at 28+ days post-dose one was 83.4% (95%CI, 54.0-94.0) (**Supplement 7**).

**Figure 1.**
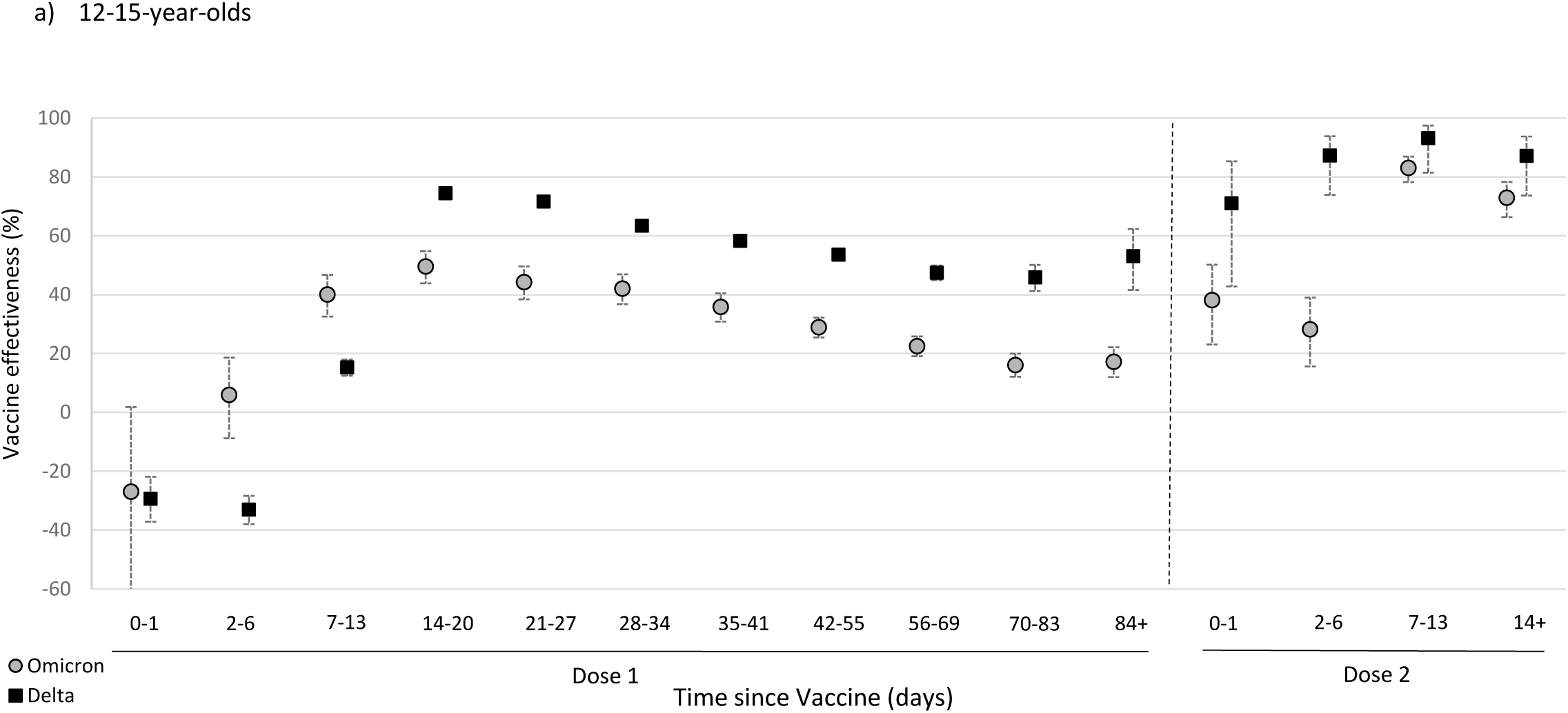
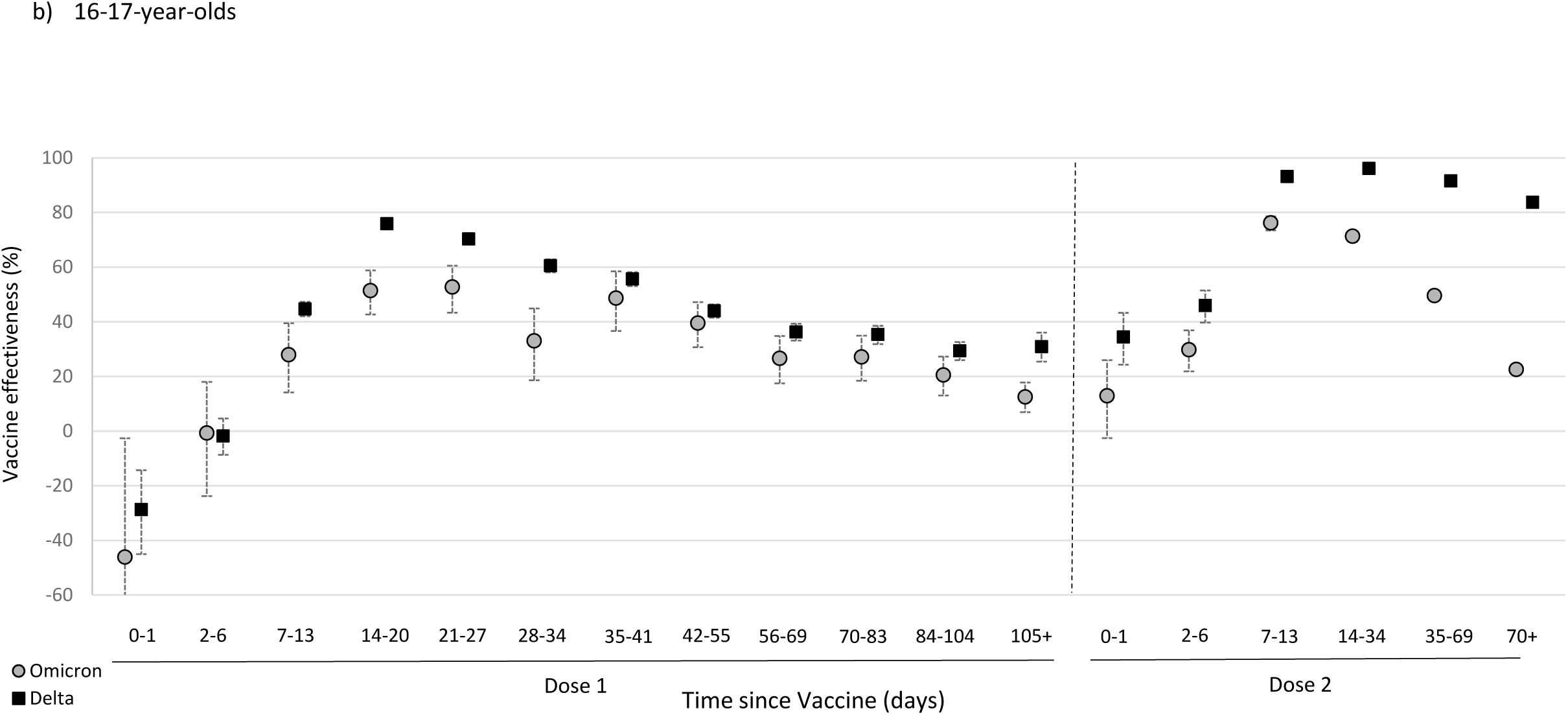
Vaccine effectiveness with 95% confidence intervals against symptomatic, PCR-confirmed COVID-19 with the Delta and Omicron variants among adolescents after one and two doses of BNT162b2 (Comirnaty, Pfizer-BioNTech) in England.

For 16-17-year-olds, VE after dose one against symptomatic disease with Delta peaked at days 14-20 (75.9%; 95%CI 74.3-77.3), declined gradually and was lowest between days 84-104 at 29.3% (95%CI 25.9-32.6) **(Figure 1, Supplement 6)**. For Omicron, the VE peak was significantly lower at 52.7% (95CI% 43.3-60.5) between days 21-27 and fell to 12.5% (95CI% 6.9-17.8) from day 105 (15 weeks) onwards. After dose two, VE peaked after 14-34 days at 96.1% (95%CI 95.2-96.8) for Delta and 7-13 days later at 76.1% (73.4-78.6) for Omicron, but fell rapidly for Omicron, reaching 22.6% (95%CI, 14.5-29.9) by 70+ days (10 weeks) compared to 83.7% (95%CI, 72.0-90.5) for Delta (**Supplement 6**). For Delta, VE against hospitalisation at 28+ days post-dose one was 76.3% (95%CI, 61.1-85.6) (**Supplement 7**). There was insufficient follow-up time for assessing hospitalisation after two doses and for Omicron for either age-group.

## Discussion

BNT162b2 vaccination in 12-15-year-olds and 16-17-year-olds was associated with lower VE against symptomatic COVID-19 caused by Omicron compared to Delta. After one dose, VE against symptomatic disease with Omicron dropped to ≤20% by 84+ days (12 weeks), highlighting the need for a second dose in both age groups. A second dose was associated with rapid increase in VE within two weeks of vaccination, with higher protection against Delta compared to Omicron. Among 16-17-year-olds who were vaccinated earlier than 12-15 year-olds and, therefore, had longer follow-up, VE against Omicron dropped rapidly compared to Delta, consistent with observed trends in UK adults receiving a similar two-dose extended BNT162b2 schedule.^10^ Similar trends will likely occur in 12-15-year-olds over time. Reassuringly, very high protection was achieved against hospitalisations due to Delta even after a single dose. Further follow-up is needed to assess protection against hospitalisation due to Omicron, any additional protection offered by the second dose and duration of protection against hospitalisation for both variants.

Pre-licensure trials reported 93% (mRNA-1273) to 100% (BNT162b2) efficacy in preventing COVID-19 among 12-15 year-olds from 7 days (BNT162b2) or 14 days (mRNA-1273) after two doses given 3-4 weeks apart, ^11,12^ but the short interval between doses prevents comparison with our cohort. Real-world VE data against two BNT162b2 doses includes a US study using a similar test-negative case-control design estimating 93% (95%CI, 83%–97%) VE against hospitalisation during June-September 2021,^13^ and another reporting 81% VE (95%CI, -55 to 90%) VE against hospitalisation in 12–15-year-olds, although this included only 45 cases,^14^ whilst early Israeli data estimated 91.5% (95%CI, 88.2%– 93.9%) VE against SARS-CoV-2 infection in 12–15-year-olds.^15^

This study is, so far, the only VE evaluation against the Omicron variant in adolescents after one and two mRNA vaccine doses. After two BNT162b2 doses given 8-12 weeks apart in UK adults, a similar high VE was observed for both Delta and Omicron at 93% and 76%, respectively ^10^ which was comparable to data from South Africa reporting VE of 70% against Omicron 2+ weeks after dose 2.^16^

The rapid waning of protection after the first and second BNT162b2 dose against symptomatic disease with Omicron, the now dominant variant in the UK and worldwide, indicates that the current adolescent immunisation programme as a stand-alone intervention is unlikely to sustain suppression of infections in the medium-to-long term. If the aim of the programme is to reduce infections, then regular boosters will likely be needed.

## Data Availability

Applications for relevant anonymised data should be submitted to the UK Health Security Office for Data Release: https://www.gov.uk/government/publications/accessing-public-health-england-data/about-the-phe-odr-and-accessing-data

## Ethics approval

UKHSA has legal permission, provided by Regulation 3 of The Health Service (Control of Patient Information) Regulations 2002, to process patient confidential information for national surveillance of communicable diseases and as such, individual patient consent is not required to access records.

## Supplement 1. Methods

### Data sources

Sources of data on vaccination status, SARS-CoV-2 testing, identification of variants, covariates included in the analyses and data linkage methods have been described previously.^8-10^ Testing data for adolescents were linked to NIMS on 18/01/2022 using combinations of the unique individual National Health Service (NHS) number, date of birth, surname, first name, and postcode using deterministic linkage – 92.5% of eligible tests from adolescents could be linked to the NIMS.

For adolescents, data on PCR-tests were extracted on 18/01/2022 containing data up to 12/01/2022 from week 37 2021 for 12-15-year-olds and week 32 2021 for 16-17-year-olds. Those vaccinated before the above periods were not included. These restrictions were done to reflect the time that vaccination was a general recommendation in these age groups.

Cases who reported symptoms and were tested in Pillar 2 were included in the analysis. Any negative tests taken within 7 days of a previous negative test, and any negative tests where symptom onset date was within the 10 days or a previous symptoms onset date for a negative test were dropped as these likely represent the same episode. Negative tests taken within 21 days of a subsequent positive test were also excluded as chances are high that these are false negatives. Positive and negative tests within 90 days of a previous positive test were also excluded; however, where participants had later positive tests within 14 days of a positive then preference was given to PCR tests and symptomatic tests. For adolescents who had more than one negative test, a maximum of two negative tests could be included per person (one from the period pre 22/11/2021 and one in the period post 22/11/2021). Data were restricted to persons who had reported symptoms and gave a symptom onset date within the 10 days before testing to account for reduced PCR sensitivity beyond this period in an infection event. Cases with booster doses or unusual vaccine schedules were excluded.

Variants were defined as previously described.^10^ In brief, cases were defined as the Delta or Omicron variant based on whole genome sequencing, genotyping or S-gene target status, with sequencing taking priority, followed by genotyping followed by SGTF status. Where subsequent positive tests within 14 days included sequencing, genotyping or SGTF information, this information was used to classify the variant. All cases prior to week 48 were defined as Delta, unless SGTF, genotyping or sequencing information confirmed otherwise. From week 48 onwards, tests were only included if they had SGTF, genotyping or sequencing information. The Delta analysis was restricted up to week 1 of 2022. Tests were defined as Omicron from week 48 onwards using SGTF, genotyping or sequencing information.

Testing data and linkage to the Emergency Care Dataset, as well as inclusion and exclusion factors for analysis has been described previously.^8^ Emergency care hospitalisation data were extracted on 18/01/2022 with cases and controls included if tested by 31/12/2021 to allow time for hospitalisation in cases which is defined as within 14 days of a positive test.

### Statistical analysis

We have previously reported the methodology for test-negative case-control analysis.^8-10^ Analysis was done by logistic regression with the PCR-test result as the dependent variable and vaccination status included as an independent variable. Vaccine effectiveness was defined as 1-odds of vaccination in cases/odds of vaccination in controls, with vaccination stratified by interval after each dose. Similar analyses were performed for hospitalisations with all cases included and cases defined as those admitted. Vaccine effectiveness was adjusted in logistic regression models for age, sex, index of multiple deprivation (quintile), ethnic group, geographic region (NHS region), period (calendar week of onset), clinical risk group status (a separate flag for those aged over and under 16), clinically extremely vulnerable (if aged 16 and above) and previous positivity. For children age was defined as at 31 August 2021.

**Supplement 2.**
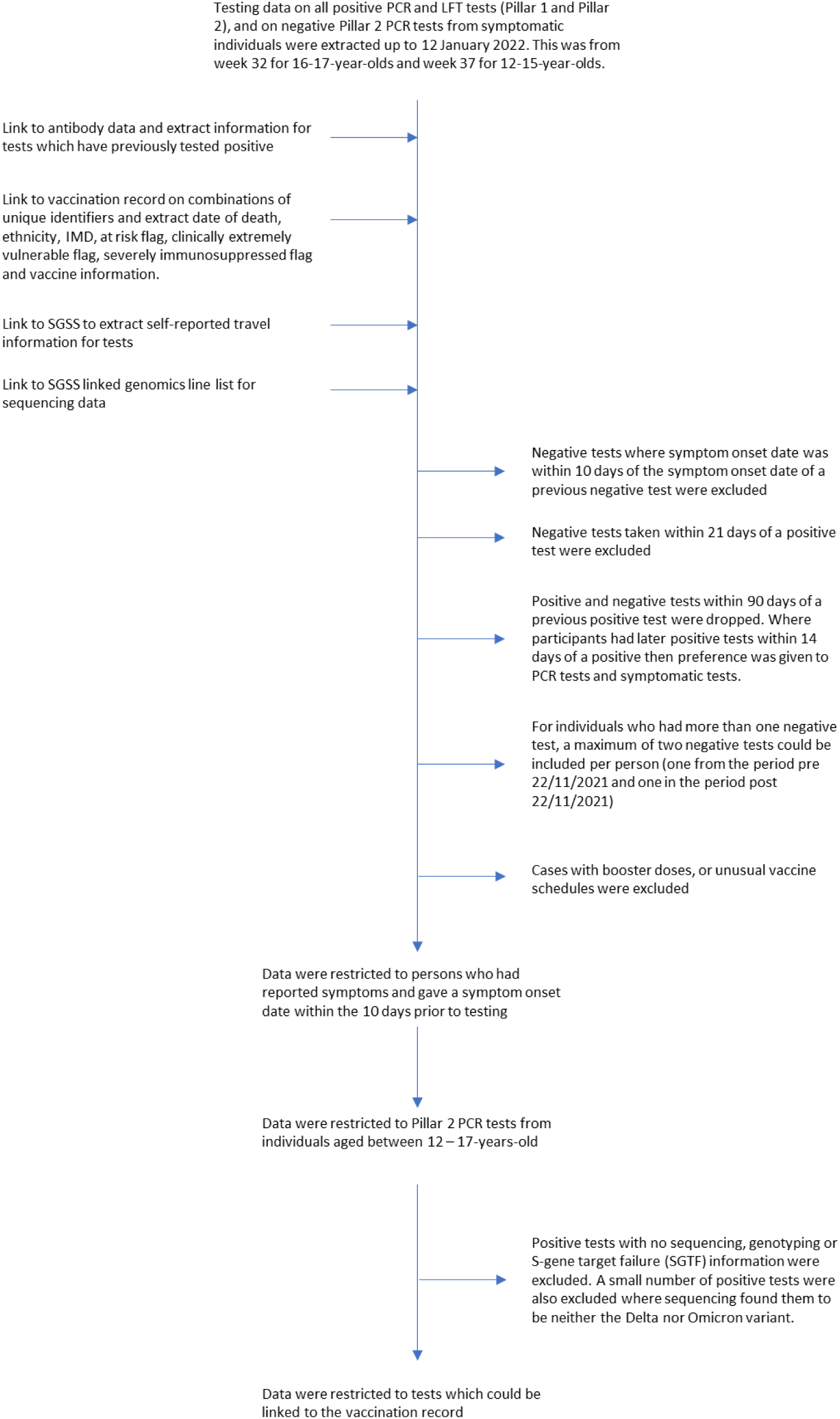
Methods Flow Chart.

**Supplement 3.**
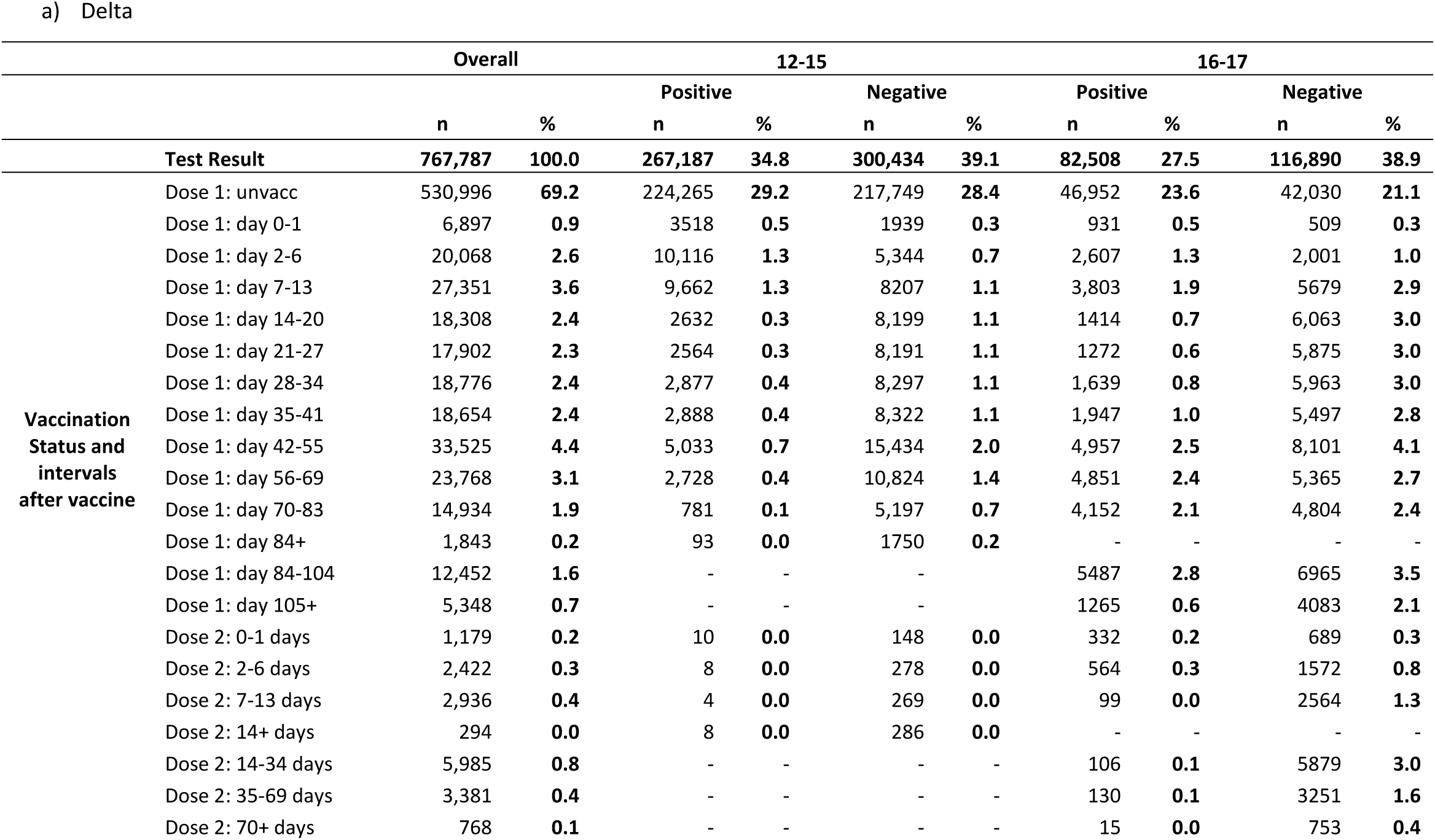
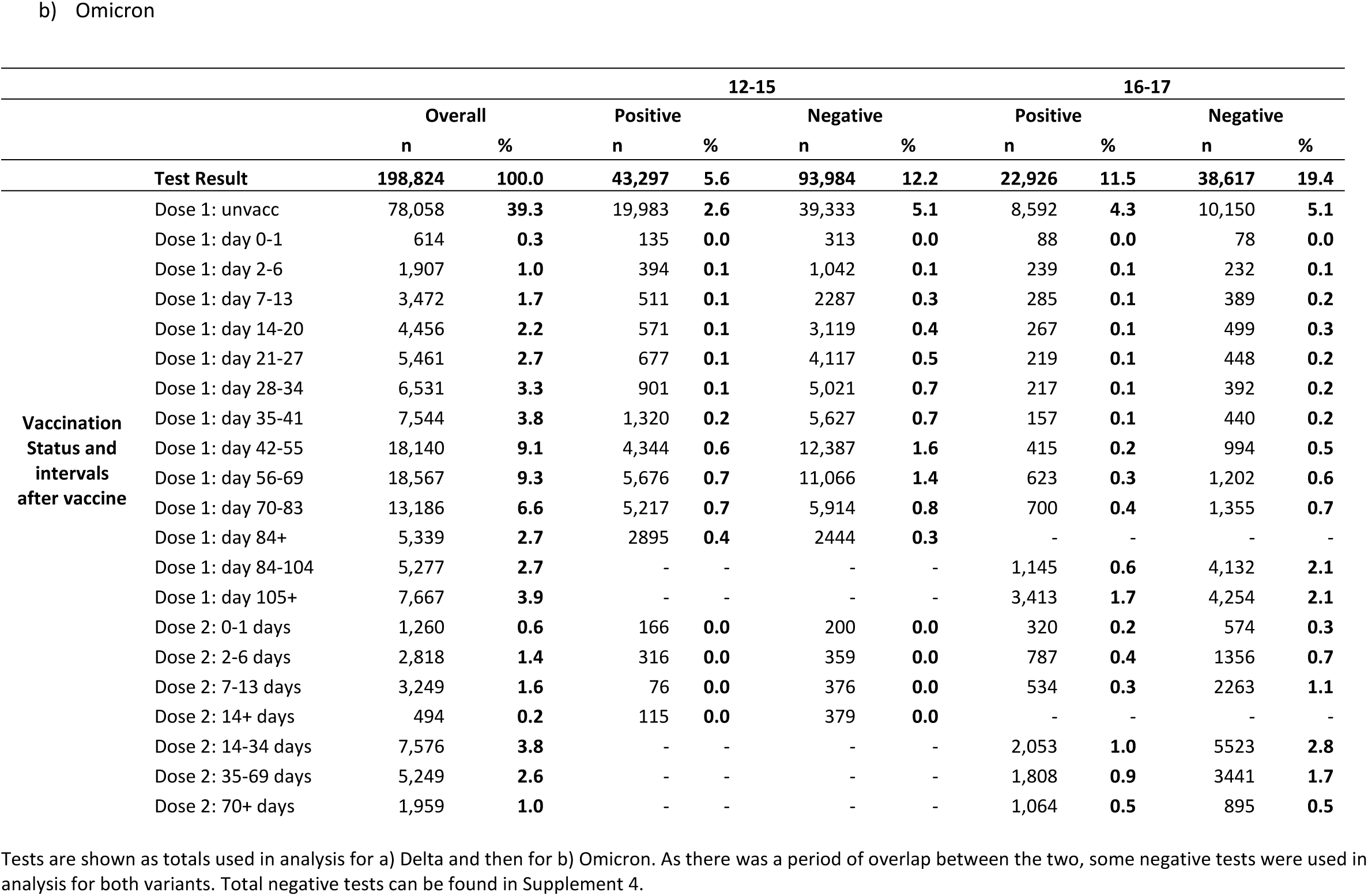
Total number of positive and negative PCR-confirmed SARS-CoV-2 test results in individuals tested for SARS-CoV-2 for grouped ages 12-15 and 16-17, in England. Data is shown for the Delta and Omicron variants.

**Supplement 4.**
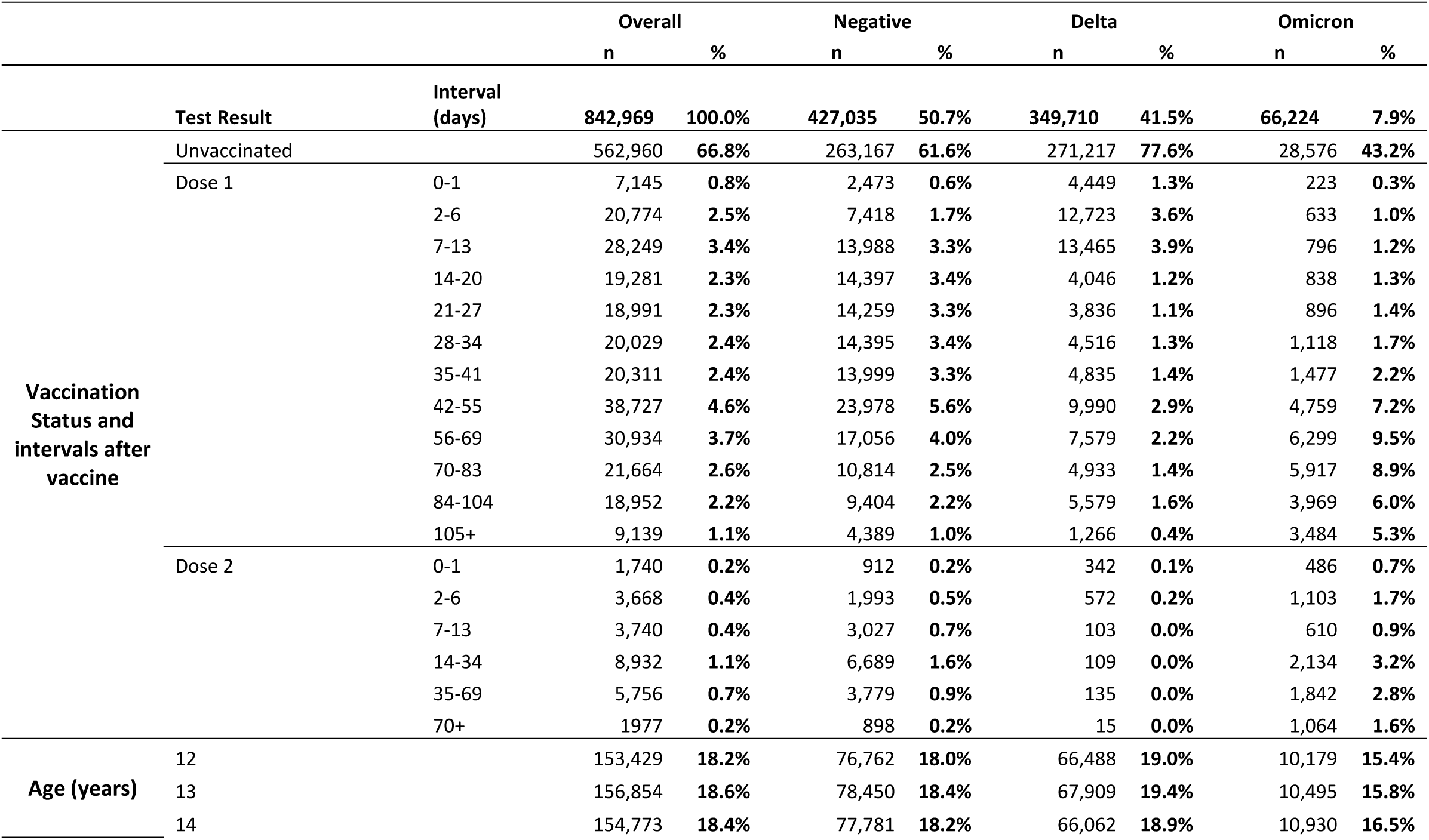
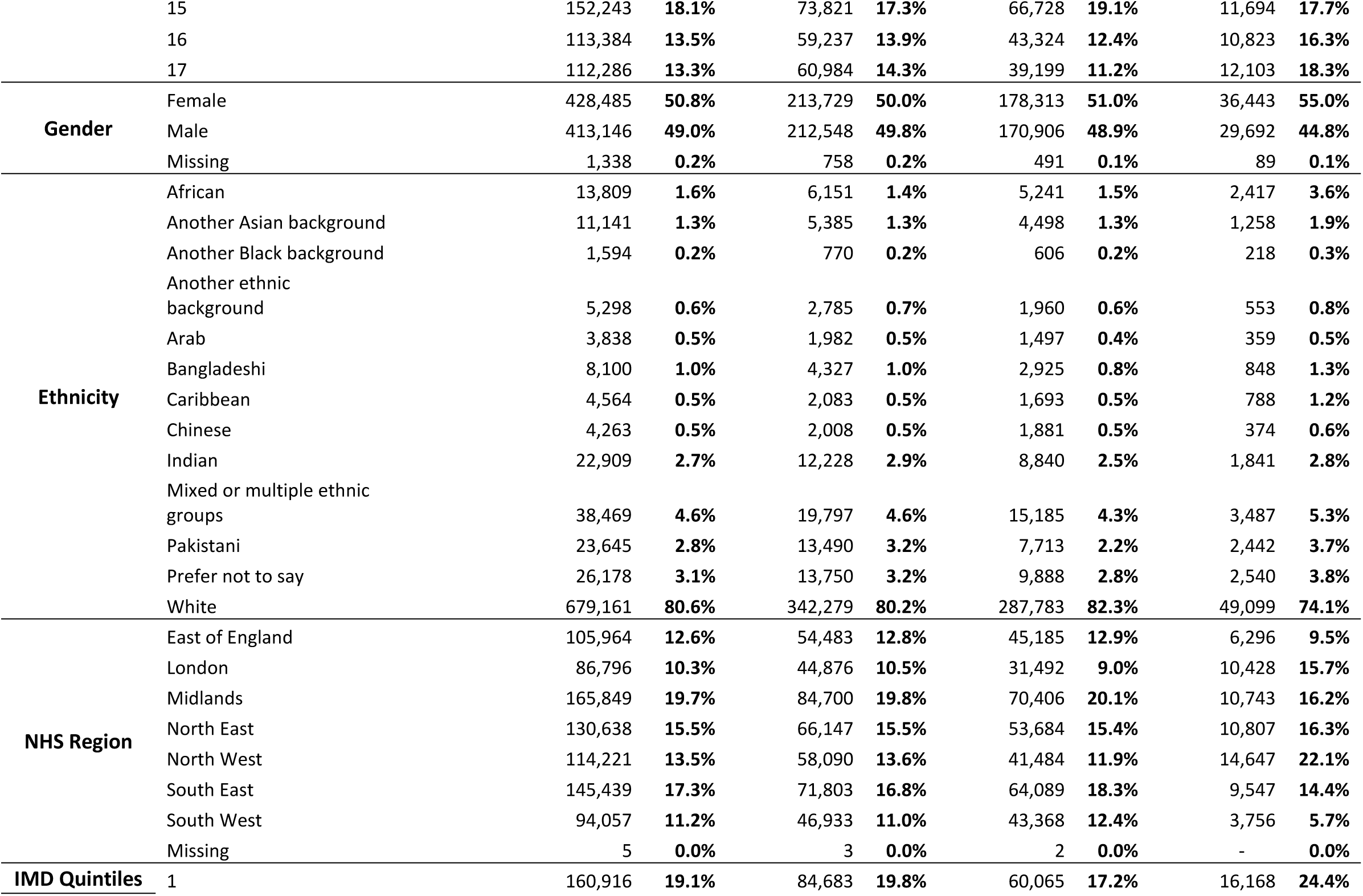
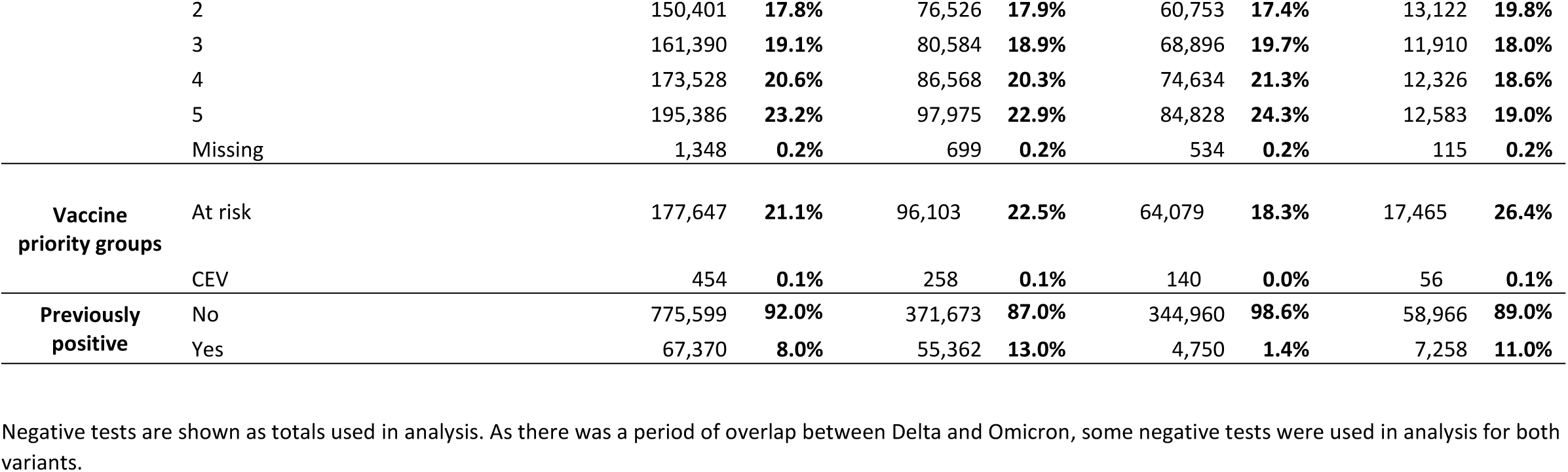
Descriptive characteristics of positive and negative PCR-confirmed SARS-CoV-2 test results among adolescents aged 12-17 years after one and two doses of BNT162b2 (Comirnaty, Pfizer-BioNTech in England. Data is shown for Delta and Omicron.

**Supplement 5.**
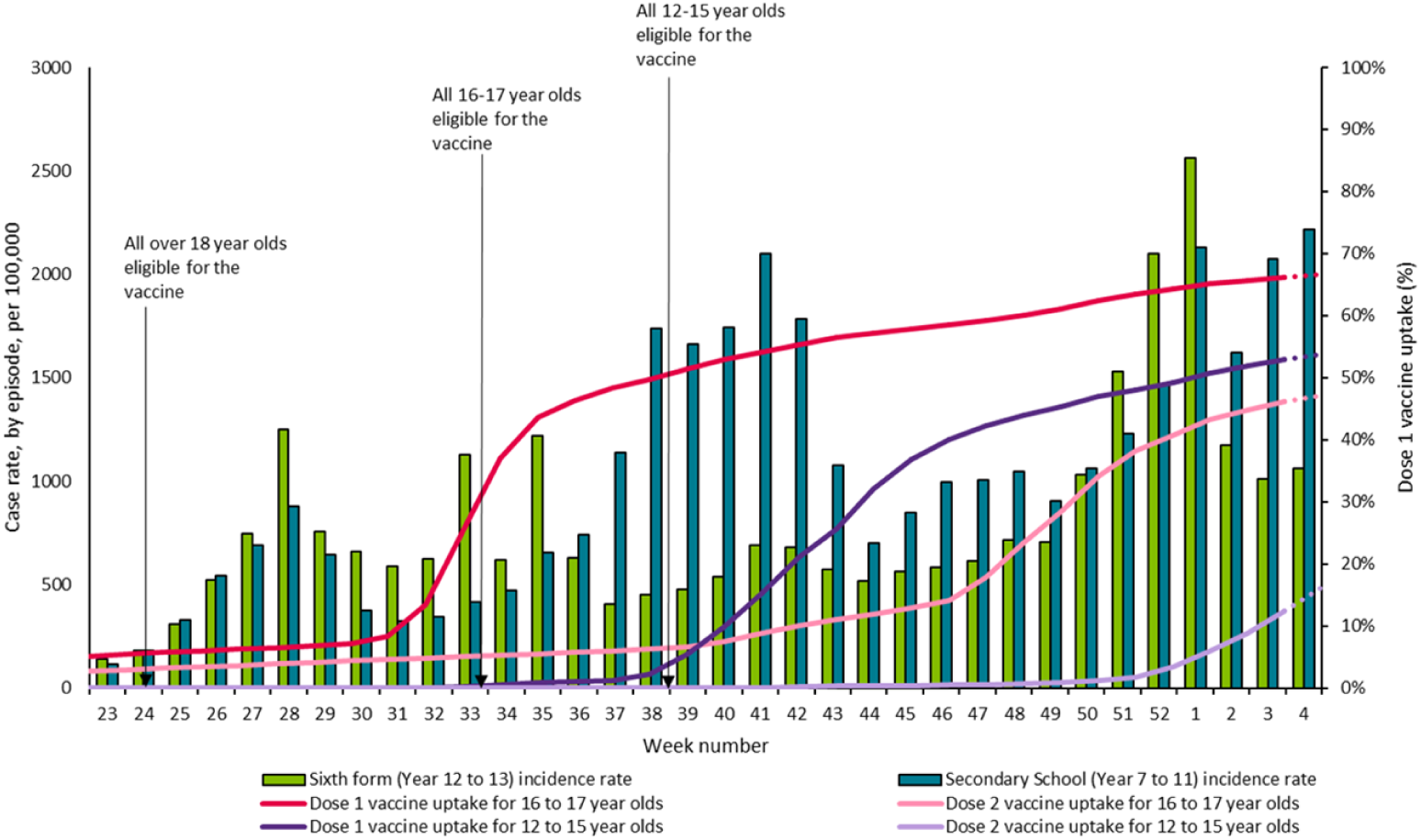
Weekly incidence of COVID-19 cases per 100,000 population from week 23 2021, in secondary age cohorts (Year 7 to 11) and sixth form age cohorts (Year 12 to Year 13) with dose 1 vaccine uptake in 12-15-year-olds and in 16-17-year-olds.

Graph taken from UKHSA Weekly Influenza and COVID-19 Surveillance graphs ^17^

Incidence definition: School age cohorts are calculated based on academic year birth cohorts. Those born between 01/09/2004 – 31/08/2005 are included in the year 12 school group and those born between 01/09/2003 – 31/08/2004 are included in the year 13 school group. Case rate denominators are sourced from ONS 2020 mid-year estimates.

Vaccine coverage definition: From Week 42 the ages are calculated based on age as of 31 August 2021. The under 50 age group includes all those aged under 50 including those born after the 31 August 2021 (denominator). Those whose date of birth is after the 31 August 2021, have an age of zero and are included in the denominator. Only vaccinations recorded as given to persons aged greater or equal to 1 have been included (numerator). Both numerators and denominators are sourced from the NIMS and exclude deaths. All data presented are for vaccinations within the living population on the date of extraction and therefore removes both formal and informal registered deaths in the numerator and denominator for the purposes of calculating vaccine uptake.

**Supplement 6.** Vaccine effectiveness with 95% confidence intervals against symptomatic, PCR-confirmed COVID-19 among adolescents and adults after one and two doses of BNT162b2 (Comirnaty, Pfizer-BioNTech) in England. Children aged 12-15 years have yet to receive their second dose of vaccine in England.

|  |  | 12-15-year-olds |  | 16-17-year-olds |  |
| --- | --- | --- | --- | --- | --- |
|  |  | Delta | Omicron | Delta | Omicron |
| Interval after vaccine |  | VE | VE | VE | VE |
| **Vaccination Status and intervals after vaccine** | Dose 1: day 0-1 | -29.3 (-37.2 to -21.9) | -26.9 (-64.1 to 1.8) | -28.8 (-45.0 to -14.3) | -46.1 (-108 to -2.6) |
|  | Dose 1: day 2-6 | -33.1 (-38.0 to -28.4) | 5.9 (-8.8 to 18.6) | -1.8 (-8.7 to 4.6) | -0.8 (-23.8 to 18.0) |
|  | Dose 1: day 7-13 | 15.3 (12.5 to 18) | 40.1 (32.6 to 46.7) | 44.7 (42.0 to 47.3) | 27.9 (14.2 to 39.5) |
|  | Dose 1: day 14-20 | 74.5 (73.2 to 75.6) | 49.6 (43.9 to 54.8) | 75.9 (74.3 to 77.3) | 51.4 (42.7 to 58.8) |
|  | Dose 1: day 21-27 | 71.7 (70.3 to 73) | 44.3 (38.4 to 49.6) | 70.3 (68.3 to 72.2) | 52.7 (43.3 to 60.5) |
|  | Dose 1: day 28-34 | 63.4 (61.7 to 65.1) | 42.1 (36.7 to 46.9) | 60.6 (58.1 to 62.9) | 33.0 (18.6 to 44.9) |
|  | Dose 1: day 35-41 | 58.4 (56.4 to 60.3) | 35.8 (30.8 to 40.5) | 55.7 (53.0 to 58.2) | 48.7 (36.6 to 58.5) |
|  | Dose 1: day 42-55 | 53.6 (51.9 to 55.2) | 28.9 (25.4 to 32.2) | 44.0 (41.5 to 46.4) | 39.5 (30.7 to 47.2) |
|  | Dose 1: day 56-69 | 47.5 (44.9 to 49.9) | 22.5 (19.1 to 25.8) | 36.3 (33.1 to 39.3) | 26.6 (17.4 to 34.8) |
|  | Dose 1: day 70-83 | 45.9 (41.2 to 50.1) | 16.1 (12.1 to 20.0) | 35.3 (31.8 to 38.6) | 27.2 (18.4 to 34.9) |
|  | Dose 1: day 84+ | 53.1 (41.6 to 62.4) | 17.2 (12.0 to 22.1) | - | - |
|  | Dose 1: day 84-104 | - | - | 29.3 (25.9 to 32.6) | 20.5 (13.0 to 27.3) |
|  | Dose 1: day 105+ | - | - | 30.9 (25.4 to 36.0) | 12.5 (6.9 to 17.8) |
|  | Dose 2: 0-1 days | 71.1 (42.8 to 85.4) | 38.1 (23.1 to 50.2) | 34.5 (24.3 to 43.3) | 12.9 (-2.6 to 26.0) |
|  | Dose 2: 2-6 days | 87.3 (73.9 to 93.8) | 28.3 (15.6 to 39.0) | 45.9 (39.7 to 51.4) | 29.7 (21.9 to 36.8) |
|  | Dose 2: 7-13 days | 93.2 (81.5 to 97.5) | 83.1 (78.2 to 86.9) | 93.1 (91.6 to 94.4) | 76.1 (73.4 to 78.6) |
|  | Dose 2: 14+ days | 87.2 (73.7 to 93.8) | 73 .0(66.4 to 78.3) | - | - |
|  | Dose 2: 14-34 days | - | - | 96.1 (95.2 to 96.8) | 71.3 (69.3 to 73.1) |
|  | Dose 2: 35-69 days | - | - | 91.5 (89.9 to 93.0) | 49.5 (45.7 to 53.0) |
|  | Dose 2: 70+ days | - | - | 83.7 (72.0 to 90.5) | 22.6 (14.5 to 29.9) |

**Supplement 7.** Vaccine effectiveness and 95% confidence intervals against hospitalisation following PCR-confirmed COVID-19 with Delta among adolescents aged 16-17 with one dose of Comirnaty (PF) in England.

a) 12-15-year-olds
|  |  | Overall |  | Positive |  | Negative |  | VE |
| --- | --- | --- | --- | --- | --- | --- | --- | --- |
|  |  | n | % | n | % | n | % |  |
| Test Result |  | 253,892 | 100.0 | 293 | 0.1 | 253,599 | 99.9 |  |
| Vaccination status and interval after vaccine | unvaccinated | 189,833 | 74.8 | 255 | 0.1 | 189,578 | 74.8 |  |
|  | d1:0-27 | 27,308 | 10.8 | 34 | 0.0 | 27,274 | 10.8 | 14.2 (-25.6 to 41.4) |
|  | d1:28+ | 36,751 | 14.5 | 4 | 0.0 | 36,747 | 14.5 | 83.4 (54.0 to 94.0) |

b) 16-17-year-olds
|  |  | Overall |  | Positive |  | Negative |  | VE |
| --- | --- | --- | --- | --- | --- | --- | --- | --- |
|  |  | n | % | n | % | n | % |  |
| Test Result |  | 86,097 | 100.0 | 171 | 0.2 | 85,926 | 99.8 |  |
| Vaccination status and interval after vaccine | unvaccinated | 34,338 | 39.9 | 130 | 0.2 | 34,208 | 39.7 |  |
|  | d1:0-27 | 17,814 | 20.7 | 19 | 0.0 | 17,795 | 20.7 | 64.6 (40.7 to 78.9) |
|  | d1:28+ | 33,945 | 39.4 | 22 | 0.0 | 33,923 | 39.4 | 76.3 (61.1 to 85.6) |

